# Genome-wide Association Clustering Meta-analysis in European and Chinese Datasets for Systemic Lupus Erythematosus identifies new genes

**DOI:** 10.1101/2023.07.07.23292357

**Authors:** Mohammad Saeed

## Abstract

Genome-wide association studies (GWAS) face multiple challenges in order to identify reliable susceptibility genes for complex disorders, such as Systemic lupus erythematosus (SLE). These include high false positivity due to number of SNPs genotyped, false negativity due to statistical corrections and the proportional signals problem. Association clustering methods, by reducing the testing burden, have increased power than single variant analysis. Here, OASIS, a locus-based test, and its novel statistic, the OASIS locus index (OLI), is applied to European (EU) and Chinese (Chi) SLE GWAS to identify common significant non-HLA, autosomal genes. Six SLE dbGAP GWAS datasets, 4 EU and 2 Chi involving 19,710 SLE cases and 30,876 controls were analyzed. OLI is defined as the product of maximum −logP at a locus with the ratio of actual to predicted number of significant SNPs and compared against the standard P-value using Box plots and Wilcoxon Signed Rank Test. OLI outperformed the standard P-value statistic in detecting true associations (Wilcoxon Signed Test Z= - 4.11, P<1×10^−4^). Top non-HLA significant loci, common in both ethnicities were 2q32.2 (STAT4, rs4274624, P=9.7×10^−66^), 1q25.3 (SMG7, rs41272536, P=3.5×10^−52^), 7q32.1 (IRF5, rs35000415, P=1.9×10^−45^), 8p23.1 (BLK, rs2736345, P=1.5×10^−25^) and 6q23.3 (TNFAIP3, rs5029937, P=4.4×10^−24^). Overall, OASIS identified 19 highly significant and 16 modestly significant (P>10^−8^) non-HLA SLE associated genes common to EU and Chi ethnicities. Interaction of these 35 genes elucidated important SLE pathways viz NOD, TLR, JAK-STAT and RIG-1. OASIS aims to advance GWAS by rapid and cost-effective identification of genes of modest significance for complex disorders.

**Key Messages:**

- GWAS are challenged by risk genes of modest effect.
- OASIS, a clustering algorithm, can help identify genes of modest significance for complex disorders such SLE, rapidly and cost-effectively using publicly available GWAS datasets.
- This meta-analysis identified 35 genes common to both European and Chinese populations.
- Interaction of these genes identify major SLE pathways to be NOD, TLR, JAK-STAT and RIG-1.

## Introduction

Systemic lupus erythematosus (SLE) is a complex disorder with significant genetic underpinnings that are only partially explained by multiple genome-wide association studies (GWAS). Identification of reliable susceptibility genes is critical for expanding the number of molecular targets for clinical testing and drug discovery. Despite the pathobiological understanding of complex disorders being greatly enhanced by GWAS, it is possible that several loci of modest significance remain to be discovered (1). This may be due to the inherent phenotypic heterogeneity of SLE, along with several other methodological challenges with GWAS.

Genotyping a multitude of single-nucleotide polymorphisms (SNPs) leads to the multiple-testing and proportional-signal problems that are the primary challenges in GWAS. The consequent high rate of false positivity prevents modest associations from being distinguished from random noise. This is dealt with by multiple testing corrections, which directly result in a high false negativity rate leading to missing heritability, a major cause of disappointment with GWAS (2). Other corrective strategies applied to tease out such modest genetic effects include GWAS meta-analyses, inclusion of different populations and biological pathway analysis (3, 4).

Another important remedial measure is association binning methods such as gene-based testing. Binning methods, by decreasing the number of tests per study, potentially increase power and reduce the multiple-testing burden. However, such tests generally work by assigning the most significant, or a weighted P-value to a gene, which potentially skew the association signal especially in high linkage disequilibrium (LD) loci. OASIS (<u>O</u>bjective <u>A</u>ssimilation of <u>S</u>NPs Interacting in <u>S</u>ynchrony) is an association clustering algorithm that takes into account all SNPs at a locus, without invoking a weighting procedure or selecting variants based on statistical association (5). By binning variants in loci, OASIS (5), provides an alternative to increasing sample size for GWAS. Moreover, OASIS merges two aspects of the LD phenomenon: strength of association (OASIS Quadrants A and B) and the number of surrounding significant SNPs (Quadrants A and C). Here, the novel OASIS Locus Index (OLI) is presented that combines the Quadrant information into a single statistic. Classically Manhattan plots are used to identify significant associations in GWAS. OLI is shown here to detect true associations at a lower significance level than the standard Manhattan plot, subverting the need for huge sample sizes. Hence, OASIS algorithm can be used to mine existing GWAS datasets for new genes of modest significance for complex disorders, such as SLE (2, 6).

Since, GWAS meta-analyses of different populations teases out common loci, OASIS was applied to six SLE dbGAP GWAS datasets, 4 Europeans (EU) (7–10) and 2 Chinese (Chi) (11, 12). This study included association statistics for 19,710 SLE cases and 30,876 controls, involving 31,736 EU and 14,159 Chi subjects. The HLA locus was excluded as its association intensity leads to the proportional signals problem (5). OASIS meta-analysis identified 183 unique loci common between the two populations. After applying appropriate corrections 35 genes were found to be significantly associated with SLE. Network analysis of these candidate genes identified several key pathobiological pathways for SLE. These findings may further unravel SLE pathogenesis and spark future clinical testing and drug discovery.

## Methods

### OASIS Algorithm

True genetic associations are accompanied by statistically significant signals from surrounding SNPs due to LD with the susceptibility variant (13). Diagrammatically, these surrounding SNPs form a cluster around the causal variant, an ‘OASIS’, observable in numerous GWAS (14–16). The OASIS algorithm has been previously described (2). Briefly, a locus is defined as a 200kb cluster (default window size that may be manually altered). P-values reported in all GWAS datasets are obtained from publically available repositories, links to which have been given in the dataset section. The first variant with a P ≤ 0.05 is considered the start of a new locus and all SNPs with P ≤ 0.05, located within 200kb of this initial SNP, are counted to define the OASIS_score_. The lowest P-value at this locus (-log.P_max_) is also reported. Expected number of significant SNPs (SNP_Expected_) is calculated from the number of genotyped SNPs (G) at the locus (G_5%_). OASIS Locus Index (OLI) is defined as −log.P_max_ x (OASIS_score_ / SNP_Expected_). Since OLI is a P-value multiplied by a ratio, it is comparable to standard P-values. Box plots and Wilcoxon Signed Rank Test is used to calculate statistical significance for OLI.

The OASIS algorithm provides an alternative to increasing sample sizes for GWAS to ascertain variants with low to moderate effect sizes. This is made possible by composite analysis based on two axes (−log.P and OASIS_score_) divided into association Quadrants A–D. This unifies two aspects of the LD phenomenon—strength of association of a single-variant and the number of significant genetic markers at a locus. The 3-sigma (3σ; three standard deviations or a value ≥99.7%of the data) cutoffs are calculated for the −log.P_max_ values and the OASIS_score_. Quadrant A loci are those that cross the 3σ cutoffs on both axes, Quadrant B loci are positive on −log.P_max_ but not on OASIS_score_ whereas, Quadrant C loci are positive on OASIS_score_ but not on −log.P_max_. Quadrant D loci fail to meet the 3σ cutoffs on either axis. By taking into account both the −log.P_max_ and the OASIS_score_, OLI converges the Quadrant data for each locus into a single statistic. Though opinion and experience exists for not using any corrections to prevent false negativity (17, 18), it was considered reasonable to use the median of OLI for Quadrant A from the combined datasets as a cutoff for evaluating the most significant loci (Figure S2).

OASIS software code in Python 2.7.9 (5) offers several features including tabulated data of 3σ significant loci with overlapping loci highlighted, links to NCBI Gene and Ensembl map as well as graphs for OLI and −log.P_max_. LD has been previously shown to maximally extend to about 2 Mb (19). Therefore, loci overlapping within 2 Mb distance were considered replicated. Moreover, this allowed a reasonable comparison between GWAS datasets, which often use different genotyping platforms over varying time periods.

OASIS is an algorithm that functions in a manner akin to global tests of association such as gene- or pathway-based tests (20, 21). Standard association analysis is based on the χ^2^ statistic, which is skewed by low-frequency alleles and can result in highly significant P-values (type I error). Clustering of significant SNPs, as in OASIS, reduces the possibility of false positive associations (type I error). The proportional signals problem, typified by the HLA-region, is dealt with by generating separate graph for −log.P_max_ > 8 (P<1×10^−8^). Replication has been the classic mechanism of verification (22). Analyzing multiple GWAS across ethnicities with OASIS allows for the robust loci to be identified by replication, even if modest in significance (4).

### Datasets

GWAS datasets were downloaded from the publicly available dbGAP and the GWAS Catalog repositories. Meta-analysis of four EU SLE datasets, phs000202 (db1)(7), phs000122 (db2)(8), GCST003156 (db3)(9) and GCST005831 (db4)(10) and two Chi SLE datasets GCST90011866 (db5)(11) and GCST90014238 (db6)(12) is conducted using OASIS. Db1 consisted of 706 SLE females and 353 controls (7), Db2 with 1435 SLE cases and 3583 controls (8), Db3 had 7,219 SLE cases and 15,991 controls (9), Db4 had 907 SLE patients and 1524 healthy controls (10). Overall, there were 10,285 EU SLE cases and 21,451 EU controls. The Chi datasets consisted of Db5 with 4222 SLE cases, 8431 controls (11) and Db6 with 512 SLE cases and 994 matched healthy controls (12). Overall, there were 4,734 Chi SLE cases and 9,425 controls. Hence, OASIS analysis was finally based on the summary statistics (SNP, location, P-values) of 19,710 SLE cases and 30,876 controls.

## Results

### OASIS Locus Index (OLI)

Across GWAS studies (db1 to db6) the number of significant SNPs (P<0.05) is directly proportional to the number of genotyped SNPs with a gradient of 0.05 (Figure S1A). This is as expected since the 5% cutoff value was set apriori for determining significance. However, there was no correlation between number of 3σ significant loci or number of significant loci in any Quadrant and number of genotyped SNPs (Figure S1B, S1C). This shows that the OASIS analysis identifying significant loci is not substantially affected by increasing number of SNPs genotyped at a locus or in a GWAS.

At each modestly significant (3σ) locus, genes are annotated by searching the most significant SNP (5×10^−2^>P>1×10^−8^) on NCBI SNP. Highly significant non-HLA loci (P<1×10^−8^) are separately analyzed by OASIS to prevent the proportional signals problem. In these loci, the SNPs were similarly annotated for their respective genes. This allowed comparison of genes identified by OASIS with the standard GWAS analysis based on highly significant P-values. Box plot analysis is carried out on genes with SNPs in at least two separate GWAS with P<1×10^−8^ and that were common to both ethnicities. The SNPs with −log.P_max_ values in a gene are selected. OLI is easily calculable for each locus as described in the methods section. Number of genotyped SNPs at each locus (G) is known and noted in the OASIS algorithm. Figure S1A forms the basis of calculating the SNP_Expected_ value as the 5% number of genotyped SNPs at a locus (G). Highest OLI for SNPs with −log.P_max_ < 8 (i.e. P>1×10^−8^) in the respective genes in concordant GWAS are noted. Box plot of this data allows comparison between OLI (limited to −log.P_max_ < 8) and the highly significant P-values (−log.P_max_ > 8) for major SLE genes across various GWAS (Figure 1). OLI outperformed standard association statistics (P-values) that are normally displayed as a Manhattan plot (Wilcoxon Signed Test Z= −4.11, P<0.0001) (Figure 1B). Low to modest effect sizes of susceptibility loci require large sample sizes. OLI is shown here to detect true associations at a lower significance level, subverting the need for huge sample sizes.

**Figure 1.**
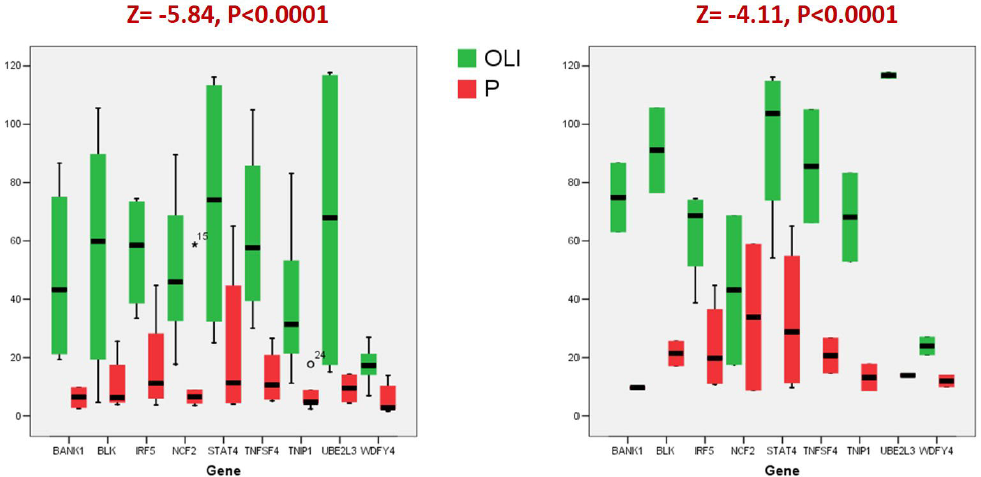
OASIS Locus Index (OLI) **A) At each gene locus −log P values for six GWAS** **B) At each locus −log P > 8 only in any GWAS** **Legend:** **OLI = −log.P_max_. (N^P<0.05^ _actual_ / N^P<0.05^ _expected_)** N^P<0.05^ is the count of significant SNPs at a given locus (OASIS) N^P<0.05^ is calculated as the 5% of genotyped SNPs at that locus OLI is restricted (by user choice) to P = 1×10^−8^ whereas the lowest P-value is unrestricted. Box plots are shown. Box Plot A shows –log.P-values for a highly significant gene locus for all six GWAS. If a locus was not highly significant (−log.P > 8) in any GWAS the highest –log.P value was chosen. Box plot B shows −log.P > 8 only i.e. highly significant P-values for major SLE genes across GWAS. OLI outperformed standard association statistics (P-values) that are normally displayed as a Manhattan plot as shown by Wilcoxon Signed Tests. Hence, this comparison shows that OLI is a more sensitive test to correctly identify a truly associated locus by detecting true associations at a lower significance level, subverting the need for huge sample sizes.

### OASIS Genome-wide Association

There were 19 highly significant (P<10^−8^) non-HLA SLE loci that replicated in both ethnicities (Table 1). Two further loci were found in only one ethnicity, ARHGAP31 in Chi and ITGAM / ITGAX in EU. STAT1-STAT4 locus had the highest significance (−log.P_max_ = 65). Other highly significant genes in both ethnicities included SMG7, IRF5, BLK and TNFAIP3 with (−log.P_max_ > 20). OASIS identified 1488 modestly significant loci (P>10^−8^), of which 775 were less than 200kb apart. These distilled to 183 unique loci that replicated in both ethnicities. Loci were included only if they replicated in at least 2 datasets of one ethnicity (e.g. EU) and 1 dataset of the other ethnicity (e.g. Chi). Median OLI of Quadrant A (OLI=50) was used as a cut off to identify robust loci in Quadrants B and C (Figure S2). All loci in Quadrant A were included in this analysis. Furthermore, type I error caused by low frequency alleles predominantly affect Quadrant B loci. OLI was less than the −log.P_max_ for 56 loci in Quadrant B, indicating lower than expected number of SNPs were significant. These loci were eliminated based on low OLI to protect against type I error. After all these corrective measures, there were 16 modestly significant loci that replicated in both EU and Chi ethnicities (Table 2). Hence, this study identified a total of 35 significant non-HLA SLE associated genes common to EU and Chi ethnicities.

**Table 1.** Highly significant (P<10^−8^) SLE loci common to EU and Chi ethnicities.

| **Locus** | **Gene** | **Hi_SNP** | **P-value** | **Max_SNP** | **P** | **OLI** | **D'** |
| --- | --- | --- | --- | --- | --- | --- | --- |
| 1p13.2 | PTPN22 | rs2476601 | 12.08 | rs115699904 | 7.17 | 70.40 | 0.968 |
| 1q23.3 | FCGR2A | rs6671847 | 11.18 | rs10800519 | 7.9 | 78.41 | 0.684 |
| 1q25.1 | TNFSF4 | rs1234313 | 13.11 | rs844644 | 7.29 | 52.54 | 0.761 |
| 1q25.3 | SMG7 | rs41272536 | 51.45 | rs12146097 | 3.95 | 20.48 | 0.756 |
| 1q25.3 | NCF2 | rs789180 | 9.37 | rs7552232 | 6.47 | 89.54 | 0.715 |
| 1q32.1 | IKBKE | rs6540432 | 8.18 | rs1930437 | 7.9 | 79.30 | 0.99 |
| 2p22.3 | RASGRP3 | rs13385731 | 17.38 | rs6746388 | 6.84 | 14.27 | 0.972 |
| 2p14 | SPRED2 | rs268124 | 8.07 | rs6740462 | 7.75 | 71.05 | 0.995 |
| 2q32.2 | STAT4 | rs4274624 | 65.01 | rs10931481 | 7.71 | 94.02 | 0.981 |
| 4q24 | BANK1 | rs10516487 | 9.79 | rs13131198 | 7.93 | 63.23 | 0.889 |
| 5q33.1 | TNIP1 | rs6889239 | 17.66 | rs960709 | 7.93 | 54.51 | 0.969 |
| 6q23.3 | TNFAIP3 | rs5029937 | 23.36 | rs657180 | 7.92 | 54.68 | 0.95 |
| 7q32.1 | IRF5 | rs35000415 | 44.73 | rs36073657 | 7.98 | 64.04 | 1 |
| 8p23.1 | BLK | rs2736345 | 24.82 | rs13277113 | 7.12 | 89.57 | 1 |
| 10q11.23 | WDFY4 | rs7097397 | 13.88 | rs1904605 | 7.85 | 86.55 | 0.911 |
| 11p15.5 | IRF7 | rs61877857 | 10.37 | rs1870727 | 7.95 | 96.83 | 0.527 |
| 16p13.13 | CLEC16A | rs12599402 | 10.99 | rs7186145 | 7.97 | 53.93 | 0.967 |
| 19p13.2 | TYK2 | rs11085727 | 12.24 | rs2304257 | 7.78 | 56.72 | 0.985 |
| 22q11.21 | UBE2L3 | rs5749502 | 13.27 | rs5754508 | 7.45 | 20.00 | 0.738 |

**Table 2.** Modestly significant (P>10^−8^) SLE loci common to EU and Chi ethnicities.

| Locus | Gene | Max_SNP E1 | P E1 | Max_SNP E2 | P E2 | OLI | D' | Q E1 | Q E2 |
| --- | --- | --- | --- | --- | --- | --- | --- | --- | --- |
| 1q25.1 | TNFSF4 | rs844642 | 5.15 | rs844644 | 7.29 | 52.54 | 0.992 | B | B |
| 3p14.3 | DNASE1L3 | rs62259783 | 7.08 | rs73081581 | 2.29 | 32.29 | 0.99 | A | C |
| 5q21.1 | ST8SIA4 | rs3846626 | 4.89 | rs1559059 | 5.85 | 57.78 | 0.834 | A | C |
| 6p21.31 | SCUBE3 | rs10755687 | 5.01 | rs2395614 | 7.97 | 108.68 | 0.993 | A | C |
| 6q23.3 | TNFAIP3 | rs657180 | 7.92 | rs657597 | 7.91 | 63.58 | 1 | B | B |
| 7q11.23 | NCF1 | rs149974255 | 4.01 | rs141214576 | 7.97 | 92.77 | 0.497 | B | B |
| 8q24.21 | PVT1 | rs12679073 | 5.50 | rs1400481 | 6.44 | 88.95 | 0.085 | A | A |
| 9p21.3 | IFNA5 | rs112280831 | 6.07 | rs10113879 | 5.00 | 80.45 | 1 | B | A |
| 11q13.1 | FOSL1 | rs12273982 | 6.61 | rs12801669 | 7.95 | 77.99 | 0.962 | A | A |
| 12p11.1 | DNM1L | rs35429841 | 7.19 | rs11059950 | 7.88 | 58.87 | 0.929 | B | A |
| 12p11.21 | DDX11 | rs139744872 | 1.90 | rs10844759 | 5.12 | 43.84 | 0.468 | C | A |
| 13q14.11 | ELF1 | rs150593918 | 3.78 | rs150103021 | 7.13 | 58.69 | 1 | C | A |
| 17p11.2 | TNFRSF13B | rs4404112 | 4.73 | rs55701306 | 7.86 | 66.59 | 0.921 | B | B |
| 17q25.1 | GRB2 | rs4789169 | 5.87 | rs59641546 | 3.77 | 52.40 | 0.956 | B | C |
| 19q13.33 | CD37 | rs10417155 | 2.96 | rs2278405 | 6.18 | 72.98 | 0.664 | C | A |
| 19q13.11 | PDCD5 | rs259265 | 3.62 | rs12461589 | 6.60 | 56.68 | 0.466 | C | A |

### SLE Pathways

In order to understand the pathways involved in SLE pathogenesis, protein interactions using STRING were performed (23). All candidate genes in SLE associated loci identified by OASIS (Table 1 and 2) that were common between EU and Chi GWAS, were evaluated. Five additional significant genes that were found in only a single ethnicity were also included. These were ARHGAP31 in Chi only and EU only genes ITGAM, ITGAX, IFIH1 and REEP1. Gene relationships that were based only on experimentation or co-expression were included. In order to allow maximum interactions to surface, low confidence (0.15) network was selected. A highly integrated network was formed with a PPI enrichment P< 1.0×10^−16^. Such an enrichment indicates that the proteins are at least partially biologically connected, as a group. The network had a total of 38 nodes involving 67 connections. Mean node degree was 3.53, indicating average number of interactions (Table S1).

The most significant pathways that emerged from this analysis (Table S2) were NOD receptor pathway involving 7 genes (OR = 1.32, P =4.24×10^−6^), TLR with 5 genes (OR=1.41, P =6.83×10^−5^), and the JAK-STAT pathway with 5 genes (OR=1.21, P =4.5 ×10^−4^). However, the pathway with the highest strength of association was the RIG-I-like receptor signaling pathway involving 4 genes (OR= 1.47 and P = 3.8×10^−4^) (Figure 2).

**Figure 2.**
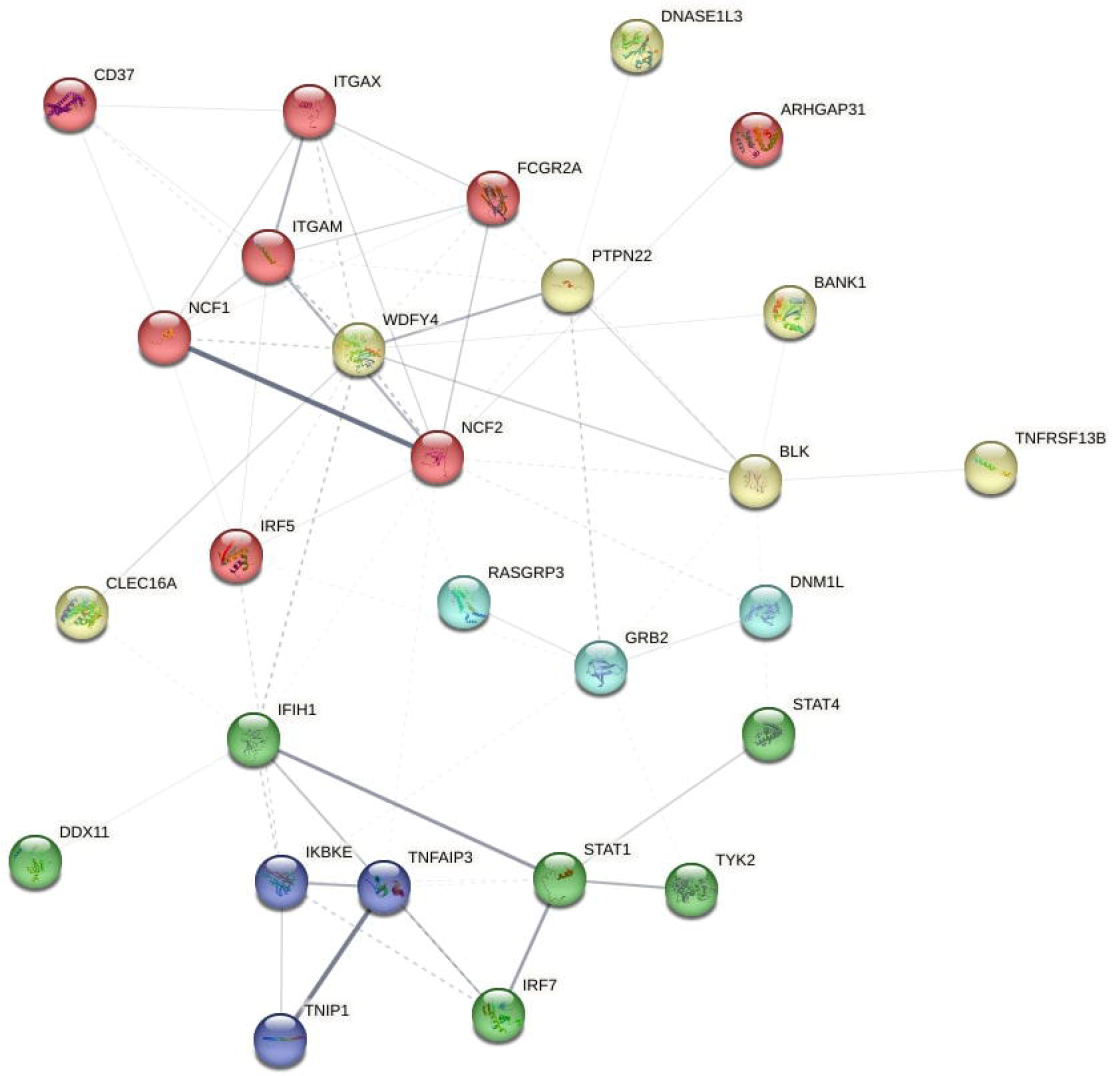
Pathway analysis was performed using STRING to detect protein interactions. **Legend:** OASIS identified 35 SLE associated genes common between EU and Chi GWAS and 5 additional significant genes in at least one ethnicity. These were evaluated for protein interactions using STRING. The network was restricted to co-expression or experimentally determined interactions. In order to allow maximum interactions to surface, low confidence (0.15) network was selected. PPI enrichment P-value was highly significant : < 1.0e-16

## Discussion

GWAS analysis face multiple challenges in order to identify reliable susceptibility genes for complex disorders. These include high false positivity driven by large number of SNPs genotyped, false negativity due to modest associations and statistical corrections and the proportional signals problem. OASIS attempts to help advance GWAS by using a clustering approach to reduce the number of tests, thereby increasing the power of the study and separately analyzing high intensity association signals (P<10^−8^). As aptly demonstrated here, OLI outperformed the standard P-value statistic in detecting true associations (Figure 1). Furthermore, by concomitantly analyzing multiple GWAS especially from diverse populations (e.g. EU and Chi), OASIS allows the more robust disease associations to surface.

Here, OASIS identified 19 highly significant and 16 modestly significant non-HLA SLE associated genes common to EU and Chi ethnicities. Several of these genes are novel (Table 1, 2). SMG7, involved in nonsense-mediated mRNA decay, was shown in a single study so far to be associated with SLE (24). It was shown that SMG7 regulates gene expression and contributes to autoantibody production in SLE (24). PVT1, a circulating lncRNA, was suppressed in serum of SLE patients (25). We had recently shown that DDX11 and DNM1L are associated with SLE and that their expression is altered in SLE monocytes (2). The B-cell associated CD37 receptor could impact SLE as it regulates apoptosis (26) and T-cell signaling (27). CD37 interacted with WDFY4, ITGAM, ITGAX and NCF1 in the STRING analysis (Table S1). Overall, interaction of the 35 genes (Table 1 and 2) elucidated important SLE pathways such as the NOD, TLR, JAK-STAT and RIG-1 (Figure 2). This study, therefore, gives impetus to further research to unravel SLE at the molecular level.

Genetic association studies began in earnest with two findings, candidate gene associations (CGA) of ACE I/D polymorphism with myocardial infarction (MI) (28) and APOE e4 with Alzheimer’s disease (AD) (29). At that time it was hoped that the era of personalized medicine had arrived and genetics was to provide the driving force for it (30). Technological advancements made large scale genotyping feasible and the first major genome-wide association study (GWAS) discovered CFH gene in age-related macular degeneration (22). However, with subsequent GWAS multiple issues came to light that prevented the initial enthusiastic expectation to be met. Despite very large sample sizes and high density marker screens in MI, ACE I/D or its locus could not be replicated in GWAS (31) and only a large focused meta-analysis could verify it (32). APOE e4 on the other hand was effectively replicated in GWAS (33). Careful evaluation of the ACE I/D data showed that the association signal had a strength of 10^−4^ (34), whereas the strength of APOE e4 was 10^−20^ (13). Similarly, mutations were later detected for familial amyotrophic lateral sclerosis (ALS) (35) in PON cluster genes that could not be verified in GWAS or meta-analysis (36, 37), in spite of showing significant associations in multiple populations initially (38, 39). This was due to the application of statistical corrections that increase false negatives when associations are of modest significance. Moreover, associations of haplotypes are often assessed at similar levels of significance as SNP genotype associations (17, 40) which further complicates statistical corrections.

Strength of association at a locus, so heavily relied on in GWAS led to false positive results, questioning the use of any particular cutoff as an indication of true association. Strong allelic (P=10^−12^) and haplotype (P=10^−23^) associations in MYH9 gene failed to identify a functional mutation (41). Wider mapping of the locus identified APOL1 with even higher allelic (P=10^−23^) and haplotype (P=10^−63^) associations that were functionally verified (17). Allelic heterogeneity is another cause of missing heritability in GWAS. This was aptly demonstrated in a unique experiment to evaluate the association signal of multiple known disease–causing mutations in a single gene (19). It was shown that overlapping mutant haplotypes result in diminishing of the association signal (19).

Questions such as false negativity, phenotypic heterogeneity and missing heritability are now taking center stage in the debate on gene hunting strategies (33, 42). Technology has reduced the costs of genotyping, generating, analyzing and storing large datasets. In the CGA era, positive associations incurred more costs due to follow-up studies. This has changed over time. Currently, false negatives incur higher costs as larger samples sizes are required in subsequent studies to detect missed associations. In this era of large freely available datasets, GWAS could serve as excellent screening tool for identifying significant loci. Genes in these loci could be filtered through with complementary data such as gene expression in disease relevant tissues and protein interactions etc., to identify functionally relevant candidate genes (genomic convergence), as multiple datasets with results pointing in the same direction is evidence of a true scientific finding (43). Final validation steps could then include independent verification of function (e.g. qPCR) (2) and re-sequencing for the true susceptibility variant(s). Hence, genetics will likely better fulfil its role of identifying pathophysiologic pathways.

This study shows that OASIS can be used to mine genes for complex disorders such as SLE using existing GWAS datasets. The OLI is a novel statistic that outperforms standard association and can help identify genes / loci of modest significance without the need and expense for huge sample sizes. OASIS clustering approach attempts to circumvent the missing heritability due to allelic heterogeneity, though this may not be comprehensively possible. Further, by analyzing multiple GWAS together from varied ethnicities, the most consistent genes can be identified. These features of OASIS are geared towards allowing genomics to spearhead biology in a cost-effective manner. At times, genomics has faltered behind pathobiology, despite its organized and unbiased approaches. TDP-43 was shown to co-localize with ubiquitinated neuronal inclusions in ALS (44) and it was not until two-years later that mutations in its coding gene, TARDBP, were identified (45). Similarly, the IL23 pathway was known to be involved in the pathogenesis of Crohn’s disease (CD) 3-years prior (46) to the identification of IL23R gene using GWAS (14). With OASIS and its novel statistic OLI, there is further hope that genomics will fulfil its long awaited mission of personalized medicine by unraveling complex disorders such as SLE, rapidly and cost-effectively. The study confirms several known SLE genes as key to its pathogenesis by verifying them in six GWAS of two major world ethnicities, i.e. EU and Chi. Moreover, the novel genes identified here (Table 1 and 2) provide targets for biological studies to confirm their pathogenic relevance in SLE. Overall, this panel of 35 genes may be investigated in clinical trials to provide diagnostic and therapeutic targets for SLE modulation.

## Web Resources

dbGAP: www.ncbi.nlm.nih.gov/gap/

GWAS Catalog: www.ebi.ac.uk/gwas/downloads/summary-statistics

## Data Availability

All data produced in the present work are contained in the manuscript or available from web resources with links in the manuscript.

## Notes

### Competing Interest Statement

The authors have declared no competing interest.

### Funding Statement

This study did not receive any funding

