## Supplementary material for "Genome-wide Association Clustering Meta-analysis in European and Chinese Datasets for Systemic Lupus Erythematosus identifies new genes": Table S1

**Table S1: STRING protein-protein interactions of OASIS identified SLE genes.**

| **Gene 1** | **Gene 2** | **Homology** | **Co-expression** | **Biological interaction** | **Combined score** |
| --- | --- | --- | --- | --- | --- |
| NCF1 | NCF2 | 0 | 0.305 | 0.979 | 0.985 |
| NCF2 | NCF1 | 0 | 0.305 | 0.979 | 0.985 |
| TNFAIP3 | TNIP1 | 0 | 0.159 | 0.884 | 0.899 |
| TNIP1 | TNFAIP3 | 0 | 0.159 | 0.884 | 0.899 |
| IFIH1 | STAT1 | 0 | 0.712 | 0 | 0.712 |
| STAT1 | IFIH1 | 0 | 0.712 | 0 | 0.712 |
| IRF7 | STAT1 | 0 | 0.653 | 0 | 0.653 |
| STAT1 | IRF7 | 0 | 0.653 | 0 | 0.653 |
| ITGAM | ITGAX | 0.962 | 0.547 | 0 | 0.547 |
| ITGAX | ITGAM | 0.962 | 0.547 | 0 | 0.547 |
| IKBKE | TNFAIP3 | 0 | 0.118 | 0.502 | 0.542 |
| TNFAIP3 | IKBKE | 0 | 0.118 | 0.502 | 0.542 |
| ITGAM | NCF2 | 0 | 0.533 | 0 | 0.533 |
| NCF2 | ITGAM | 0 | 0.533 | 0 | 0.533 |
| PTPN22 | WDFY4 | 0 | 0.532 | 0 | 0.532 |
| WDFY4 | PTPN22 | 0 | 0.532 | 0 | 0.532 |
| STAT1 | TYK2 | 0 | 0.062 | 0.518 | 0.528 |
| TYK2 | STAT1 | 0 | 0.062 | 0.518 | 0.528 |
| NCF2 | WDFY4 | 0 | 0.431 | 0 | 0.431 |
| WDFY4 | NCF2 | 0 | 0.431 | 0 | 0.431 |
| ITGAM | WDFY4 | 0 | 0.429 | 0 | 0.429 |
| WDFY4 | ITGAM | 0 | 0.429 | 0 | 0.429 |
| IFIH1 | IRF7 | 0 | 0.423 | 0 | 0.423 |
| IRF7 | IFIH1 | 0 | 0.423 | 0 | 0.423 |
| IFIH1 | WDFY4 | 0 | 0.4 | 0 | 0.4 |
| WDFY4 | IFIH1 | 0 | 0.4 | 0 | 0.4 |
| FCGR2A | NCF2 | 0 | 0.366 | 0 | 0.366 |
| NCF2 | FCGR2A | 0 | 0.366 | 0 | 0.366 |
| BLK | WDFY4 | 0 | 0.325 | 0.089 | 0.359 |
| WDFY4 | BLK | 0 | 0.325 | 0.089 | 0.359 |
| GRB2 | PTPN22 | 0 | 0.054 | 0.346 | 0.355 |
| IFIH1 | IKBKE | 0 | 0.116 | 0.3 | 0.355 |
| IKBKE | IFIH1 | 0 | 0.116 | 0.3 | 0.355 |
| PTPN22 | GRB2 | 0 | 0.054 | 0.346 | 0.355 |
| STAT1 | STAT4 | 0.945 | 0.056 | 0.345 | 0.355 |
| STAT4 | STAT1 | 0.945 | 0.056 | 0.345 | 0.355 |
| IKBKE | IRF7 | 0 | 0.12 | 0.295 | 0.354 |
| IRF7 | IKBKE | 0 | 0.12 | 0.295 | 0.354 |
| NCF1 | WDFY4 | 0 | 0.352 | 0 | 0.352 |
| WDFY4 | NCF1 | 0 | 0.352 | 0 | 0.352 |
| ITGAX | WDFY4 | 0 | 0.338 | 0 | 0.338 |
| WDFY4 | ITGAX | 0 | 0.338 | 0 | 0.338 |
| IKBKE | TNIP1 | 0 | 0.11 | 0.281 | 0.333 |
| TNIP1 | IKBKE | 0 | 0.11 | 0.281 | 0.333 |
| IRF7 | TNFAIP3 | 0 | 0.111 | 0.27 | 0.323 |
| TNFAIP3 | IRF7 | 0 | 0.111 | 0.27 | 0.323 |
| FCGR2A | ITGAX | 0 | 0.313 | 0 | 0.313 |
| ITGAX | FCGR2A | 0 | 0.313 | 0 | 0.313 |
| ITGAX | NCF2 | 0 | 0.306 | 0 | 0.306 |
| NCF2 | ITGAX | 0 | 0.306 | 0 | 0.306 |
| FCGR2A | ITGAM | 0 | 0.304 | 0 | 0.304 |
| ITGAM | FCGR2A | 0 | 0.304 | 0 | 0.304 |
| CLEC16A | WDFY4 | 0 | 0.302 | 0 | 0.302 |
| WDFY4 | CLEC16A | 0 | 0.302 | 0 | 0.302 |
| ITGAM | NCF1 | 0 | 0.296 | 0 | 0.296 |
| NCF1 | ITGAM | 0 | 0.296 | 0 | 0.296 |
| ITGAX | NCF1 | 0 | 0.295 | 0 | 0.295 |
| NCF1 | ITGAX | 0 | 0.295 | 0 | 0.295 |
| BLK | PTPN22 | 0 | 0.26 | 0.078 | 0.289 |
| PTPN22 | BLK | 0 | 0.26 | 0.078 | 0.289 |
| IKBKE | IRF5 | 0 | 0.096 | 0.231 | 0.276 |
| IRF5 | IKBKE | 0 | 0.096 | 0.231 | 0.276 |
| GRB2 | RASGRP3 | 0 | 0 | 0.249 | 0.248 |
| RASGRP3 | GRB2 | 0 | 0 | 0.249 | 0.248 |
| CD37 | WDFY4 | 0 | 0.231 | 0 | 0.231 |
| DNM1L | GRB2 | 0 | 0.062 | 0.214 | 0.231 |
| GRB2 | DNM1L | 0 | 0.062 | 0.214 | 0.231 |
| WDFY4 | CD37 | 0 | 0.231 | 0 | 0.231 |
| IRF5 | WDFY4 | 0 | 0.219 | 0 | 0.219 |
| WDFY4 | IRF5 | 0 | 0.219 | 0 | 0.219 |
| DNM1L | NCF2 | 0 | 0 | 0.217 | 0.217 |
| NCF2 | DNM1L | 0 | 0 | 0.217 | 0.217 |
| BLK | FCGR2A | 0 | 0 | 0.213 | 0.213 |
| FCGR2A | BLK | 0 | 0 | 0.213 | 0.213 |
| FCGR2A | WDFY4 | 0 | 0.211 | 0 | 0.211 |
| WDFY4 | FCGR2A | 0 | 0.211 | 0 | 0.211 |
| ARHGAP31 | NCF2 | 0 | 0.116 | 0.141 | 0.208 |
| NCF2 | ARHGAP31 | 0 | 0.116 | 0.141 | 0.208 |
| BLK | NCF2 | 0 | 0.187 | 0.056 | 0.2 |
| IRF5 | NCF2 | 0 | 0.2 | 0 | 0.2 |
| NCF2 | BLK | 0 | 0.187 | 0.056 | 0.2 |
| NCF2 | IRF5 | 0 | 0.2 | 0 | 0.2 |
| CD37 | ITGAX | 0 | 0.184 | 0.058 | 0.199 |
| ITGAX | CD37 | 0 | 0.184 | 0.058 | 0.199 |
| CD37 | NCF1 | 0 | 0.197 | 0 | 0.197 |
| NCF1 | CD37 | 0 | 0.197 | 0 | 0.197 |
| IRF5 | ITGAM | 0 | 0.193 | 0 | 0.193 |
| ITGAM | IRF5 | 0 | 0.193 | 0 | 0.193 |
| BLK | TNFRSF13B | 0 | 0.179 | 0.057 | 0.192 |
| TNFRSF13B | BLK | 0 | 0.179 | 0.057 | 0.192 |
| BANK1 | WDFY4 | 0 | 0.184 | 0 | 0.184 |
| ITGAM | PTPN22 | 0 | 0.133 | 0.098 | 0.184 |
| PTPN22 | ITGAM | 0 | 0.133 | 0.098 | 0.184 |
| WDFY4 | BANK1 | 0 | 0.184 | 0 | 0.184 |
| NCF2 | PTPN22 | 0 | 0.182 | 0 | 0.182 |
| PTPN22 | NCF2 | 0 | 0.182 | 0 | 0.182 |
| DDX11 | IFIH1 | 0 | 0.065 | 0.159 | 0.18 |
| IFIH1 | DDX11 | 0 | 0.065 | 0.159 | 0.18 |
| IKBKE | STAT1 | 0 | 0.085 | 0.14 | 0.179 |
| STAT1 | IKBKE | 0 | 0.085 | 0.14 | 0.179 |
| CD37 | ITGAM | 0 | 0.163 | 0.058 | 0.178 |
| ITGAM | CD37 | 0 | 0.163 | 0.058 | 0.178 |
| GRB2 | IKBKE | 0 | 0.052 | 0.156 | 0.166 |
| IKBKE | GRB2 | 0 | 0.052 | 0.156 | 0.166 |
| IRF5 | NCF1 | 0 | 0.165 | 0 | 0.165 |
| NCF1 | IRF5 | 0 | 0.165 | 0 | 0.165 |
| STAT1 | TNFAIP3 | 0 | 0.082 | 0.128 | 0.165 |
| TNFAIP3 | STAT1 | 0 | 0.082 | 0.128 | 0.165 |
| FCGR2A | NCF1 | 0 | 0.163 | 0 | 0.163 |
| NCF1 | FCGR2A | 0 | 0.163 | 0 | 0.163 |
| CLEC16A | IFIH1 | 0 | 0.162 | 0 | 0.162 |
| IFIH1 | CLEC16A | 0 | 0.162 | 0 | 0.162 |
| DNASE1L3 | PTPN22 | 0 | 0.161 | 0 | 0.161 |
| PTPN22 | DNASE1L3 | 0 | 0.161 | 0 | 0.161 |
| IFIH1 | TNFAIP3 | 0 | 0.159 | 0 | 0.159 |
| TNFAIP3 | IFIH1 | 0 | 0.159 | 0 | 0.159 |
| BANK1 | BLK | 0 | 0.124 | 0.078 | 0.158 |
| BLK | STAT4 | 0 | 0.062 | 0.14 | 0.158 |
| BLK | BANK1 | 0 | 0.124 | 0.078 | 0.158 |
| STAT4 | BLK | 0 | 0.062 | 0.14 | 0.158 |
| RASGRP3 | WDFY4 | 0 | 0.157 | 0 | 0.157 |
| WDFY4 | RASGRP3 | 0 | 0.157 | 0 | 0.157 |
| GRB2 | IRF5 | 0 | 0 | 0.157 | 0.156 |
| IRF5 | GRB2 | 0 | 0 | 0.157 | 0.156 |
| BLK | GRB2 | 0.713 | 0 | 0.156 | 0.155 |
| GRB2 | BLK | 0.713 | 0 | 0.156 | 0.155 |
| GRB2 | TYK2 | 0 | 0 | 0.156 | 0.155 |
| TYK2 | GRB2 | 0 | 0 | 0.156 | 0.155 |
| NCF2 | TNFAIP3 | 0 | 0.155 | 0 | 0.154 |
| TNFAIP3 | NCF2 | 0 | 0.155 | 0 | 0.154 |
| IFIH1 | NCF2 | 0 | 0.153 | 0 | 0.152 |
| ITGAX | PTPN22 | 0 | 0.099 | 0.098 | 0.152 |
| NCF2 | IFIH1 | 0 | 0.153 | 0 | 0.152 |
| PTPN22 | ITGAX | 0 | 0.099 | 0.098 | 0.152 |
