## Supplementary material for "Genome-wide Association Clustering Meta-analysis in European and Chinese Datasets for Systemic Lupus Erythematosus identifies new genes": Table S2

**Table S2: STRING pathways of OASIS identified SLE genes common in EU and Chi GWAS.**

| **KEGG Pathway #** | **KEGG Pathway Description** | **Gene count** | **Strength** | **FDR** | **Gene network** |
| --- | --- | --- | --- | --- | --- |
| hsa04621 | NOD-like receptor signaling pathway | 7 | 1.32 | 4.24E-06 | STAT1,IRF7,TYK2,DNM1L,IKBKE,TNFAIP3,IFNA5 |
| hsa04620 | Toll-like receptor signaling pathway | 5 | 1.41 | 6.83E-05 | IRF5,STAT1,IRF7,IKBKE,IFNA5 |
| hsa04622 | RIG-I-like receptor signaling pathway | 4 | 1.47 | 0.00038 | IFIH1,IRF7,IKBKE,IFNA5 |
| hsa04630 | JAK-STAT signaling pathway | 5 | 1.21 | 0.00045 | STAT1,STAT4,GRB2,TYK2,IFNA5 |
| hsa04145 | Phagosome | 4 | 1.16 | 0.004 | FCGR2A,NCF1,NCF2,ITGAM |
| hsa04623 | Cytosolic DNA-sensing pathway | 3 | 1.4 | 0.0057 | IRF7,IKBKE,IFNA5 |
| hsa04658 | Th1 and Th2 cell differentiation | 3 | 1.25 | 0.0124 | STAT1,STAT4,TYK2 |
| hsa04657 | IL-17 signaling pathway | 3 | 1.23 | 0.0138 | FOSL1,IKBKE,TNFAIP3 |
| hsa04670 | Leukocyte transendothelial migration | 3 | 1.15 | 0.0212 | NCF1,NCF2,ITGAM |
