## Supplementary material for "Genome-wide Association Clustering Meta-analysis in European and Chinese Datasets for Systemic Lupus Erythematosus identifies new genes": Figure S1

**Figure S1: OASIS statistics based on genotyped SNPs**

A) The number of significant SNPs (P<0.05) is directly proportional to genotyped SNPs. Across all six GWAS studies (db1 to db6) the gradient is on average 0.05, as set apriori

Number of significant SNPs (P<0.05)

| Number of genotyped SNPs | db1 |
| --- | --- |
| 599513 | 44056 |
| 655022 | 50246 |
| 471676 | 32935 |
| 570791 | 37191 |
| 510369 | 35364 |
| 535078 | 30974 |
| 457007 | 30295 |
| 436586 | 34488 |
| 338647 | 20480 |
| 400739 | 26642 |
| 389925 | 28664 |
| 378928 | 25842 |
| 292785 | 19863 |
| 256590 | 19919 |
| 212842 | 14402 |
| 237713 | 18240 |
| 202437 | 18369 |
| 224047 | 14720 |
| 173712 | 14204 |
| 174584 | 11273 |
| 107592 | 6332 |
| 107442 | 8985 |

| Number of genotyped SNPs | db2 |
| --- | --- |
| 692194 | 42293 |
| 771227 | 45395 |
| 572705 | 32316 |
| 668131 | 38503 |
| 600771 | 33825 |
| 639436 | 30100 |
| 521529 | 30274 |
| 521919 | 31044 |
| 394093 | 19528 |
| 470683 | 26831 |
| 452572 | 26244 |
| 429736 | 22991 |
| 346231 | 19416 |
| 300683 | 20101 |
| 243169 | 14741 |
| 249591 | 15139 |
| 213041 | 16050 |
| 256362 | 13070 |
| 162531 | 9968 |
| 191566 | 9932 |
| 119976 | 7689 |
| 111279 | 5752 |

| Number of genotyped SNPs | db3 |
| --- | --- |
| 37531 | 1902 |
| 40583 | 2133 |
| 33864 | 1802 |
| 29973 | 1600 |
| 31150 | 1820 |
| 30760 | 1669 |
| 27018 | 1576 |
| 28517 | 1921 |
| 24161 | 1333 |
| 26051 | 1442 |
| 24325 | 1384 |
| 24304 | 1494 |
| 18764 | 1055 |
| 16659 | 890 |
| 14943 | 831 |
| 15142 | 846 |
| 12854 | 717 |
| 15179 | 865 |
| 8209 | 474 |
| 12801 | 686 |
| 7468 | 425 |
| 7357 | 368 |

| Number of genotyped SNPs | db4 |
| --- | --- |
| 19671 | 1058 |
| 21141 | 1168 |
| 18137 | 933 |
| 15927 | 829 |
| 16227 | 866 |
| 16179 | 878 |
| 13802 | 692 |
| 15434 | 871 |
| 13313 | 709 |
| 13070 | 738 |
| 12203 | 671 |
| 12699 | 708 |
| 9684 | 526 |
| 8349 | 552 |
| 7565 | 397 |
| 7510 | 415 |
| 6951 | 438 |
| 8937 | 527 |
| 4658 | 300 |
| 6614 | 345 |
| 4644 | 224 |
| 4428 | 297 |

| Number of genotyped SNPs | db5 |
| --- | --- |
| 575922 | 27195 |
| 566514 | 29457 |
| 494731 | 22095 |
| 494692 | 24405 |
| 432602 | 18514 |
| 496296 | 19592 |
| 390269 | 19284 |
| 272174 | 11622 |
| 219477 | 10448 |
| 348773 | 15507 |
| 344727 | 17338 |
| 334426 | 14315 |
| 273246 | 12935 |
| 247801 | 12623 |
| 213789 | 10283 |
| 216330 | 10269 |
| 184895 | 9070 |
| 210840 | 9800 |
| 159137 | 8498 |
| 154287 | 7026 |
| 100967 | 4515 |
| 94931 | 3881 |

| Number of genotyped SNPs | db6 |
| --- | --- |
| 449648 | 33207 |
| 485485 | 32486 |
| 429900 | 25454 |
| 436628 | 22073 |
| 371082 | 22458 |
| 417199 | 26950 |
| 344958 | 19857 |
| 279209 | 18065 |
| 219886 | 14833 |
| 304330 | 20002 |
| 294461 | 20260 |
| 279592 | 18104 |
| 220175 | 12260 |
| 191862 | 11057 |
| 162945 | 10213 |
| 159216 | 12507 |
| 136940 | 10653 |
| 161950 | 9594 |
| 112324 | 8735 |
| 116673 | 7231 |
| 78770 | 4893 |
| 65904 | 6351 |

B) Number of 3σ significant loci is not correlated with genotyped SNPs as shown here from data in db1. However, as shown above, number of significant SNPs is directly correlated with genotyped SNPs.

Number of SNPs (P<0.05) or OASIS Loci (>3s)

| Number of Genotyped SNPS | Sig_SNPs | Sig_Loci |
| --- | --- | --- |
| 599513 | 44056 | 958 |
| 655022 | 50246 | 1013 |
| 471676 | 32935 | 827 |
| 570791 | 37191 | 809 |
| 510369 | 35364 | 763 |
| 535078 | 30974 | 674 |
| 457007 | 30295 | 659 |
| 436586 | 34488 | 604 |
| 338647 | 20480 | 471 |
| 400739 | 26642 | 556 |
| 389925 | 28664 | 565 |
| 378928 | 25842 | 572 |
| 292785 | 19863 | 411 |
| 256590 | 19919 | 379 |
| 212842 | 14402 | 325 |
| 237713 | 18240 | 336 |
| 202437 | 18369 | 333 |
| 224047 | 14720 | 325 |
| 173712 | 14204 | 248 |
| 174584 | 11273 | 255 |
| 107592 | 6332 | 147 |
| 107442 | 8985 | 150 |

C) Number of significant loci in any OASIS Quadrant does not correlate with number of genotyped SNPs as shown from db1 data. OASIS significant loci are not affected by increasing number of SNPs genotyped at a locus or in a GWAS.

3s significant loci in Quadrants

| Number of Genotyped SNPs | Q_A | Q_B | Q_C |
| --- | --- | --- | --- |
| 599513 | 1 | 16 | 18 |
| 655022 | 7 | 7 | 16 |
| 471676 | 2 | 9 | 3 |
| 570791 | 2 | 1 | 11 |
| 510369 | 2 | 9 | 9 |
| 535078 | 2 | 4 | 8 |
| 457007 | 5 | 8 | 13 |
| 436586 | 14 | 4 | 13 |
| 338647 | 0 | 2 | 2 |
| 400739 | 2 | 2 | 9 |
| 389925 | 4 | 4 | 9 |
| 378928 | 1 | 9 | 3 |
| 292785 | 1 | 5 | 7 |
| 256590 | 0 | 0 | 7 |
| 212842 | 0 | 0 | 2 |
| 237713 | 5 | 6 | 5 |
| 202437 | 3 | 7 | 5 |
| 224047 | 1 | 1 | 2 |
| 173712 | 1 | 7 | 7 |
| 174584 | 0 | 1 | 1 |
| 107592 | 0 | 0 | 2 |
| 107442 | 1 | 2 | 1 |
| 692194 | 0 | 9 | 13 |
| 771227 | 0 | 16 | 12 |
| 572705 | 1 | 7 | 9 |
| 668131 | 3 | 3 | 15 |
| 600771 | 0 | 9 | 16 |
| 639436 | 1 | 10 | 12 |
| 521529 | 2 | 9 | 9 |
| 521919 | 1 | 10 | 18 |
| 394093 | 1 | 9 | 6 |
| 470683 | 2 | 8 | 6 |
| 452572 | 1 | 7 | 12 |
| 429736 | 1 | 1 | 3 |
| 346231 | 1 | 3 | 3 |
| 300683 | 0 | 6 | 11 |
| 243169 | 0 | 1 | 2 |
| 249591 | 3 | 3 | 3 |
| 213041 | 2 | 7 | 5 |
| 256362 | 0 | 3 | 1 |
| 162531 | 1 | 1 | 5 |
| 191566 | 0 | 2 | 1 |
| 119976 | 0 | 0 | 5 |
| 111279 | 0 | 0 | 2 |
| 37531 | 0 | 6 | 2 |
| 40583 | 3 | 6 | 5 |
| 33864 | 0 | 5 | 6 |
| 29973 | 0 | 3 | 3 |
| 31150 | 1 | 5 | 2 |
| 30760 | 0 | 3 | 5 |
| 27018 | 1 | 4 | 2 |
| 28517 | 10 | 4 | 10 |
| 24161 | 1 | 2 | 3 |
| 26051 | 0 | 3 | 1 |
| 24325 | 1 | 4 | 3 |
| 24304 | 1 | 6 | 4 |
| 18764 | 2 | 0 | 2 |
| 16659 | 1 | 1 | 4 |
| 14943 | 0 | 1 | 1 |
| 15142 | 1 | 2 | 3 |
| 12854 | 1 | 3 | 0 |
| 15179 | 0 | 3 | 1 |
| 8209 | 0 | 1 | 0 |
| 12801 | 0 | 3 | 1 |
| 7468 | 0 | 0 | 2 |
| 7357 | 0 | 0 | 1 |
| 19671 | 0 | 6 | 3 |
| 21141 | 0 | 4 | 4 |
| 18137 | 0 | 9 | 3 |
| 15927 | 0 | 3 | 2 |
| 16227 | 0 | 3 | 5 |
| 16179 | 0 | 1 | 2 |
| 13802 | 0 | 10 | 4 |
| 15434 | 0 | 2 | 7 |
| 13313 | 0 | 4 | 7 |
| 13070 | 3 | 4 | 3 |
| 12203 | 0 | 5 | 3 |
| 12699 | 0 | 4 | 5 |
| 9684 | 0 | 1 | 2 |
| 8349 | 1 | 5 | 6 |
| 7565 | 0 | 1 | 1 |
| 7510 | 0 | 4 | 2 |
| 6951 | 0 | 4 | 3 |
| 8937 | 1 | 2 | 5 |
| 4658 | 0 | 1 | 1 |
| 6614 | 2 | 2 | 1 |
| 4644 | 1 | 2 | 2 |
| 4428 | 0 | 3 | 3 |
| 575922 | 2 | 7 | 13 |
| 566514 | 1 | 17 | 14 |
| 494731 | 0 | 3 | 13 |
| 494692 | 1 | 7 | 18 |
| 432602 | 0 | 4 | 9 |
| 496296 | 3 | 3 | 13 |
| 390269 | 0 | 7 | 12 |
| 272174 | 2 | 5 | 4 |
| 219477 | 1 | 1 | 3 |
| 348773 | 0 | 8 | 7 |
| 344727 | 2 | 6 | 10 |
| 334426 | 0 | 5 | 8 |
| 273246 | 0 | 2 | 5 |
| 247801 | 0 | 4 | 6 |
| 213789 | 0 | 11 | 3 |
| 216330 | 1 | 5 | 6 |
| 184895 | 0 | 2 | 1 |
| 210840 | 0 | 2 | 6 |
| 159137 | 2 | 4 | 5 |
| 154287 | 0 | 4 | 2 |
| 100967 | 0 | 2 | 3 |
| 94931 | 0 | 3 | 1 |
| 449648 | 4 | 19 | 11 |
| 485485 | 4 | 9 | 15 |
| 429900 | 0 | 6 | 13 |
| 436628 | 1 | 3 | 3 |
| 371082 | 2 | 3 | 5 |
| 417199 | 2 | 8 | 17 |
| 344958 | 0 | 6 | 7 |
| 279209 | 2 | 4 | 6 |
| 219886 | 2 | 2 | 5 |
| 304330 | 0 | 5 | 7 |
| 294461 | 6 | 8 | 6 |
| 279592 | 2 | 7 | 5 |
| 220175 | 3 | 0 | 1 |
| 191862 | 1 | 2 | 2 |
| 162945 | 0 | 5 | 2 |
| 159216 | 2 | 3 | 5 |
| 136940 | 1 | 4 | 4 |
| 161950 | 0 | 1 | 3 |
| 112324 | 2 | 3 | 5 |
| 116673 | 0 | 0 | 1 |
| 78770 | 0 | 2 | 1 |
| 65904 | 0 | 1 | 5 |
