## Supplementary material for "Genome-wide Association Clustering Meta-analysis in European and Chinese Datasets for Systemic Lupus Erythematosus identifies new genes": Figure S2

**Figure S2: OASIS Locus Index statistical correction for identifying significant loci**

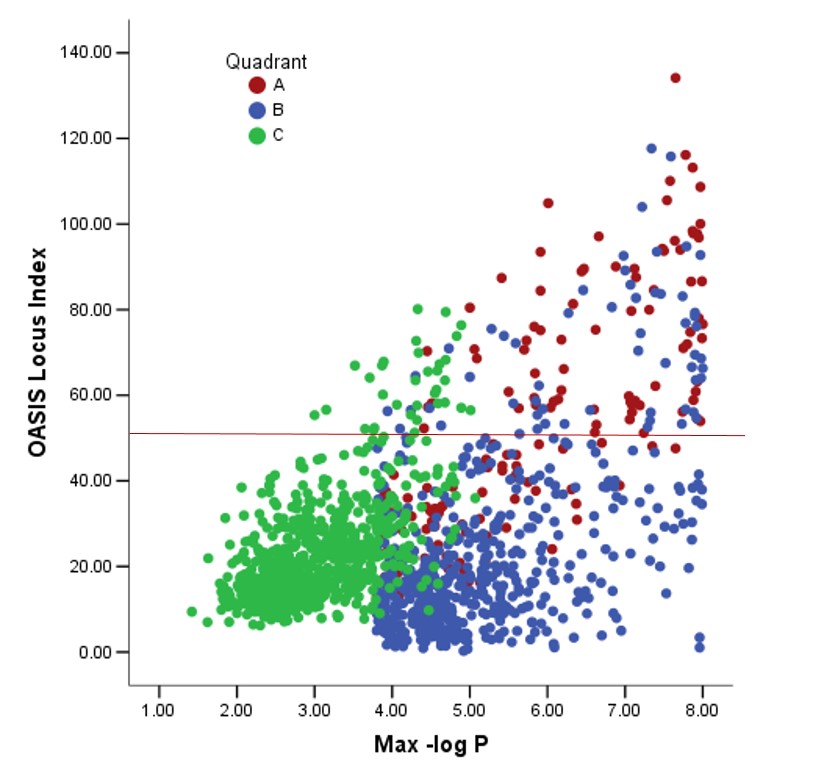

OLI was plotted against maximum –log-P values in each locus from the combined dataset of six GWAS. Each locus was significant in one of the three OASIS Quadrants (A, B or C). The median for Quadrant A was 51.3 and the mean ± SD was 54.4±26.7. For Quadrant B and C the medians were 16.6 and 19.9 respectively. Overall, the median for all three Quadrants was 20.3. Since Quadrant A, captures the most significant loci, OLI of 50 was chosen as the corrected cut off for significance of a locus.
